# Recapturing the Initial Clinical Benefit of TMS in a Major Depressive Episode: a Retrospective Analysis in a Veteran Cohort

**DOI:** 10.1101/2023.09.11.23295376

**Authors:** Jeremy Laufer, Alisa Olmsted, Irina Sampair, Michelle Madore, Jong Yoon, Laura Hack, Corey J. Keller

## Abstract

**Background:** Repetitive transcranial magnetic stimulation (TMS) is now widely accepted as an effective non-pharmacologic treatment for treatment-resistant depression. However, whether repeated acute TMS courses can recapture the antidepressant effects of the initial acute course is still an open question, especially in the Veteran population. We present here a retrospective analysis of a specialty clinic within the Veteran Affairs Hospital System to help address this question.

**Aims:** Following an acute treatment course of TMS, we sought to determine the treatment response of a subsequent TMS course. We hypothesized that those who responded to an initial acute TMS course would respond in a similar manner to a subsequent treatment course.

**Methods:** 116 cases referred for evaluation for TMS between September 2017 to April 2021 were reviewed. 63 Veterans completed at least one acute course of TMS and 12 completed at least two courses and met inclusion criteria for this review. Symptoms were evaluated via self-reported scales at baseline and weekly throughout treatment. Clinical response to subsequent treatment (>50% symptom reduction as measured by the PHQ-9) was compared to initial treatment response.

**Results:** Of the initial treatment responders (n = 6), all six responded to a second acute course, with an 85.3% symptom reduction. Of the initial treatment nonresponders (n = 6), three responded to a second acute course. Exploratory regression analysis predicted change in depression symptoms (PHQ-9) during a second TMS course using initial treatment response, time into treatment, and baseline symptom severity. Together, these factors explained 72% of the variance. No adverse events were reported in those who completed a second course, and the Veterans tolerated the treatment well.

**Conclusions:** Our findings support the growing understanding that a second acute TMS treatment course for treatment-resistant depression is safe, well-tolerated, and effective in initial responders and some non-responders. Despite multiple confounders in a naturalistic setting, robust initial treatment response was sustained in a second acute course. Low power limits generalizability, and larger powered, prospective studies are needed.

## Introduction

Major depressive disorder (MDD) is a chronic and disabling condition with a lifetime prevalence in the US population of 24.9% (Hasin et al., 2018). Treatment-resistant depression (TRD), defined as episodes of depression that do not respond to two or more therapies of adequate dose and duration, has been estimated to affect approximately 35% of those diagnosed with MDD (Nemeroff, 2007). FDA-approved in 2008, transcranial magnetic stimulation (TMS) is a form of brain stimulation targeting TRD. TMS involves using an electromagnetic coil to administer pulses of electromagnetic waves repetitively to stimulate cortical neurons and alter neural excitability. In the Veteran population, an acute course (4-6 weeks) of TMS has been shown to be clinically effective for treating MDD (Madore et al., 2022). Despite the effectiveness of the initial course, however, there is currently a lack of evidence to guide the delivery of subsequent TMS upon patient relapse. The Veterans Health Administration provides unique insight into the value of a second course of TMS as insurance typically does not limit patients from receiving subsequent TMS treatment courses following the initial acute phase of treatment. ‘Preservation TMS,’ was recently coined by Wilson and colleagues and is defined as TMS treatment to sustain the clinical response to a prior course of treatment (Wilson et al., 2022). Preservation TMS is an umbrella term that includes continuation, maintenance, relapse prevention, or rescue TMS. The goal of this study is to focus on a form of preservation TMS, specifically the clinical effects of a second, repeated acute course of TMS on Veterans who either relapsed or did not respond to an initial acute course of treatment.

Unfortunately, relapse rates following an acute course of TMS are high. A meta-analysis performed in 2018 found that only 43.6% of initial TMS responders maintained their antidepressant response after 12 months (Senova et al., 2019). At least one-third of patients require repeat TMS treatments in the year following successful acute treatment (Philip et al., 2016). Currently, a limited literature supports the effectiveness of repeat acute TMS courses in treating depression relapse following an initial acute course (Demirtas-Tatlidede et al., 2008; Janicak et al., 2010; Philip et al., 2016). For example, in one study, 38 of the 99 (38.4%) patients experienced symptom worsening following a positive response to an initial acute phase of TMS. When a second course was administered, 32 of the 38 (84.2%) patients reachieved symptomatic benefit (Janicak et al., 2010). While promising, these studies do not focus on Veterans, who suffer from medical and psychiatric comorbidities that complicate their clinical presentation and treatment response (Mi et al., 2017).

Here, through a secondary analysis within a large quality improvement study (Madore et al, 2021), we evaluated the clinical effectiveness of a second course of acute TMS in a Veteran cohort. To our knowledge, this is the first retrospective analysis to investigate the efficacy of a repeat acute TMS course in the Veteran population. Our central hypothesis was that those who clinically responded to their first acute course of TMS would likewise benefit from subsequent acute courses of TMS treatment. The data reported in this article provide evidence of the effectiveness of repeated courses of TMS in Veterans.

## Methods

This is a secondary analysis within a large quality improvement study to study the clinical effect of TMS in Veterans (Madore et al., 2021). Veterans who met criteria for analysis received at least two acute courses of TMS between September 2017 and April 2021. Treatments occurred at the Precision Neuromodulation Clinic (PNC), a specialty clinic located within the VA Palo Alto Healthcare System that administers TMS for Veterans with treatment-resistant depression. During an initial consultation, a psychiatrist evaluated TMS eligibility as determined by clinical guidelines to confirm diagnosis of a major depressive episode and medication resistance, and assess for contraindications. Individuals recommended for TMS presented to the clinic for measurement of motor threshold (MT) and for TMS treatment. All Veterans provided written consent for TMS treatment. After consultation and consent, Veterans were typically administered 30 total TMS treatment sessions, as is standard in the field (McClintock et al., 2018), with some variability based on time of clinical response and tolerability.

### Patients

Included in this analysis were all Veterans who met diagnostic criteria for MDD and completed at least two acute courses of TMS treatment. Veterans with a primary diagnosis of bipolar disorder or borderline personality disorder who received treatment were excluded from the analysis. Following an initial acute TMS treatment course, veterans needed to have completed a minimum of 15 treatment sessions in a subsequent treatment course separated from the initial course by at least 30 days. The purpose of this was to exclude situations of continuation TMS. Veterans were also excluded if weekly PHQ-9 assessments were not available for the majority of the treatment course or if clinical scores prior to the initial treatment course were low (PHQ-9 Score ≤ 5), indicating no-to-minimal depression at the beginning of treatment.

A total of 116 Veterans that received a TMS consultation were reviewed. 63 Veterans received at least one acute course of TMS treatment. Of these, 15 Veterans completed at least two courses of TMS treatment. Two Veterans with bipolar disorder were excluded. One Veteran with borderline personality disorder was excluded. Demographic information is summarized in *Table 1*. The sample was predominantly male, over the age of 50, and all were retired or unemployed. Veterans reported a recurrent history of MDD for many years with multiple psychiatric comorbidities. 42% reported a history of a least one suicide attempt. No adverse events were reported in either group during the second course.

**Table 1.** Baseline characteristics including demographic features and pertinent clinical indicators. *M, mean; SD, standard deviation; n, number of individuals; MDD, Major Depressive Disorder; PTSD, Post-Traumatic Stress Disorder*.

|  | **TMS Treatment Groups** |  |
| --- | --- | --- |
|  | Initial Responders<br>(n=6) | Initial Non-Responders<br>(n=6) |
| **Demographic Features** |  |  |
| Age in years, M(SD) | 57.5 (21.0) | 52.2 (8.4) |
| Biological Sex, n(%) |  |  |
| Male | 7 (100%) | 5 (83%) |
| Female | 0 (0%) | 1 (17%) |
| Ethnicity, n(%) |  |  |
| Caucasian | 4 (67%) | 3 (50%) |
| African American | 0 (0%) | 2 (33%) |
| Latin American | 1 (17%) | 0 (0%) |
| Asian American | 0 (0%) | 1 (17%) |
| Other | 1 (17%) | 0 (0%) |
| Marital Status, n(%) |  |  |
| Married | 4 (67%) | 4 (67%) |
| Divorced | 0 (0%) | 1 (17%) |
| Never Married | 2 (33%) | 1 (17%) |
| Education, n(%) |  |  |
| Some College | 3 (50%) | 2 (33%) |
| Degree | 3 (50%) | 4 (67%) |
| Occupation, n(%) |  |  |
| Retired | 3 (50%) | 1 (17%) |
| Unemployed | 3 (50%) | 5 (83%) |
| **Clinical Indicators** |  |  |
| Years since diagnosis, M(SD) | 20 (18.1) | 14.8 (12.7) |
| Secondary Diagnoses, n(%) |  |  |
| Anxiety Disorder | 1 (17%) | 3 (50%) |
| PTSD | 5 (83%) | 3 (50%) |
| Substance Use Disorder | 2 (33%) | 3 (50%) |
| Alcohol Use Disorder | 2 (33%) | 3 (50%) |
| Stimulant Use Disorder | 1 (17%) | 1 (17%) |
| Neurocognitive Disorder | 1 (17%) | 0 (0%) |
| Sleep Disorder | 1 (17%) | 1 (17%) |
| Suicide Attempts, n(%) |  |  |
| No | 5 (83%) | 3 (50%) |
| Yes | 2 (33%) | 3 (50%) |
| Number of Attempts, M | 2.5 | 2.3 |

### Treatment Parameters and Device

Veterans received TMS via the MagStim Horizon or Rapid2 with a figure-of-8 coil. Head dimensions were measured to localize the approximate area on the motor cortex controlling the right thumb using the Beam F3 method (Mir-Moghtadaei et al., 2015). Motor threshold was identified using the PEST program and reported as a percentage of maximal machine output (Borckardt et al., 2006). 10 out of 12 Veterans received 10Hz or intermittent theta burst stimulation (iTBS) stimulation to the left dorsolateral prefrontal cortex (DLPFC), targeted at 120% of resting motor threshold, adjusted for tolerability. Two Veterans received 1Hz TMS to the right DLPFC with one veteran receiving stimulation at 120% of MT and the other 90% of maximal machine output because a motor threshold could not be determined. Typical treatment was typically once daily, but near the latter part of the review period, Veterans also were offered to opt for multiple iTBS sessions in a single day given new clinical evidence (Cole et al., 2022, 2020). PHQ-9 scores were used to group Veterans into initial treatment responders (>50% reduction of scores after treatment) and nonresponders (<50% reduction of scores after treatment). Treatment regimens and pre- and post-treatment PHQ-9 scores are summarized in *Table 2*.

**Table 2.** Treatment parameters, time elapsed between treatment courses, and PHQ-9 scores. *PHQ-9, Patient Health Questionnaire-9; M, mean; SD, standard deviation; n, number of individuals;*

|  | TMS Treatment Groups |  |
| --- | --- | --- |
|  | Initial Responders<br>(n=6) | Initial Non-Responders<br>(n=6) |
| **Initial Course** |  |  |
| Treatment Regimen, n(%) |  |  |
| 10 Hz | 2 (33%) | 3 (50%) |
| 1 Hz | 1 (17%) | 1 (17%) |
| iTBS | 3 (50%) | 2 (33%) |
| Treatment Target, n(%) |  |  |
| Left DLPFC | 5 (83%) | 5 (83%) |
| Right DLPFC | 1 (17%) | 1 (17%) |
| Treatment Intensity, M(SD) |  |  |
| % Machine Output | 65% (12%) | 73% (15%) |
| % Motor Threshold | 120% (0%) | 113% (13%) |
| Total Sessions, M(SD) | 26.9 (6.9) | 29.7 (2.4) |
| **Second Course** |  |  |
| Treatment Regimen, n(%) |  |  |
| 10 Hz | 3 (50%) | 1 (17%) |
| 1 Hz | 1 (17%) | 1 (17%) |
| iTBS | 2 (33%) | 4 (67%) |
| Treatment Target, n(%) |  |  |
| Left DLPFC | 5 (83%) | 5 (83%) |
| Right DLPFC | 1 (17%) | 1 (17%) |
| Treatment Intensity, M(SD) |  |  |
| % Machine Output | 66.8 (12.5) | 75 (11) |
| % Motor Threshold | 120 (0) | 118 (4) |
| Total Sessions, M(SD) | 23.5 (6.9) | 32.3 (7.8) |
| **Time elapsed until second course, M(SD)** | 7.0 mo (5.8) | 16 mo (10.8) |
| **PHQ-9 Score, M(SD)** |  |  |
| Baseline, initial | 13.2 (4.7) | 20.5 (5.3) |
| Post-Treatment, initial | 2.0 (2.0) | 17.8 (6.2) |

### Clinical Evaluations

Initial symptom severity was measured at baseline prior to each TMS treatment course as well as every calendar week throughout the course. At each timepoint, Veterans completed the Patient Health Questionnaire (PHQ)-9 (Kroenke et al., 2001), and a subset of Veterans also completed the Brief Symptom Inventory-18 (BSI-18) (Derogatis, 2001).

### Statistical Analysis

The primary objective of this study was to determine if and how clinical treatment response to a subsequent acute TMS course is impacted by the initial TMS treatment course outcome. The primary outcome measure was PHQ-9 change from baseline to post-treatment during the second course of TMS. Participants were categorized into treatment responders or non-responders based on their change in PHQ-9 during the initial treatment course. In alignment with standard practice, a 50% or greater decrease in PHQ-9 score was defined as ‘treatment response,’ and PHQ-9 < 5 was defined as ‘remission.’ Missing post-treatment scores were imputed by last values carried forward (Coley et al., 2020). Student t-tests were performed to assess differences in demographics and baseline scores on clinical surveys between treatment responders and non-responders. Repeated measures ANOVA was performed to assess differences in clinical response between courses. A significance level of p ≤ 0.05 was used for all tests of significance.

To explore the relationship between symptom severity, time, and prior treatment response, a multiple linear regression analysis was performed. The target variable was PHQ-9 score in the second course. Predictor variables were either: 1) disease severity, quantified by pre-treatment PHQ-9 score, 2) time, indicated by the number of weeks into treatment, and 3) initial treatment response, calculated by the difference in pre- and post-treatment PHQ-9 in the initial course. We used backward factor selection to control for collinearity with a factor removal criterion of a partial F-statistic with p < .05. Regression residuals autocorrelation was tested with the Durbin-Watson statistic. Statistical analyses were completed using RStudio Version 3.6.1 and IBM SPSS Statistics version 28.

## Results

A first TMS treatment course improved clinical symptoms of our cohort (n=12) by an average of 49.2% (PHQ reduction). In a second treatment course, TMS improved symptoms by 63.7%. This difference was not significant (Fig 1A; Δ PHQ-9 1.8 ± 7.8, p=0.44; 14.5%Δ ± 30.7, p=0.34).

**Figure 1.**
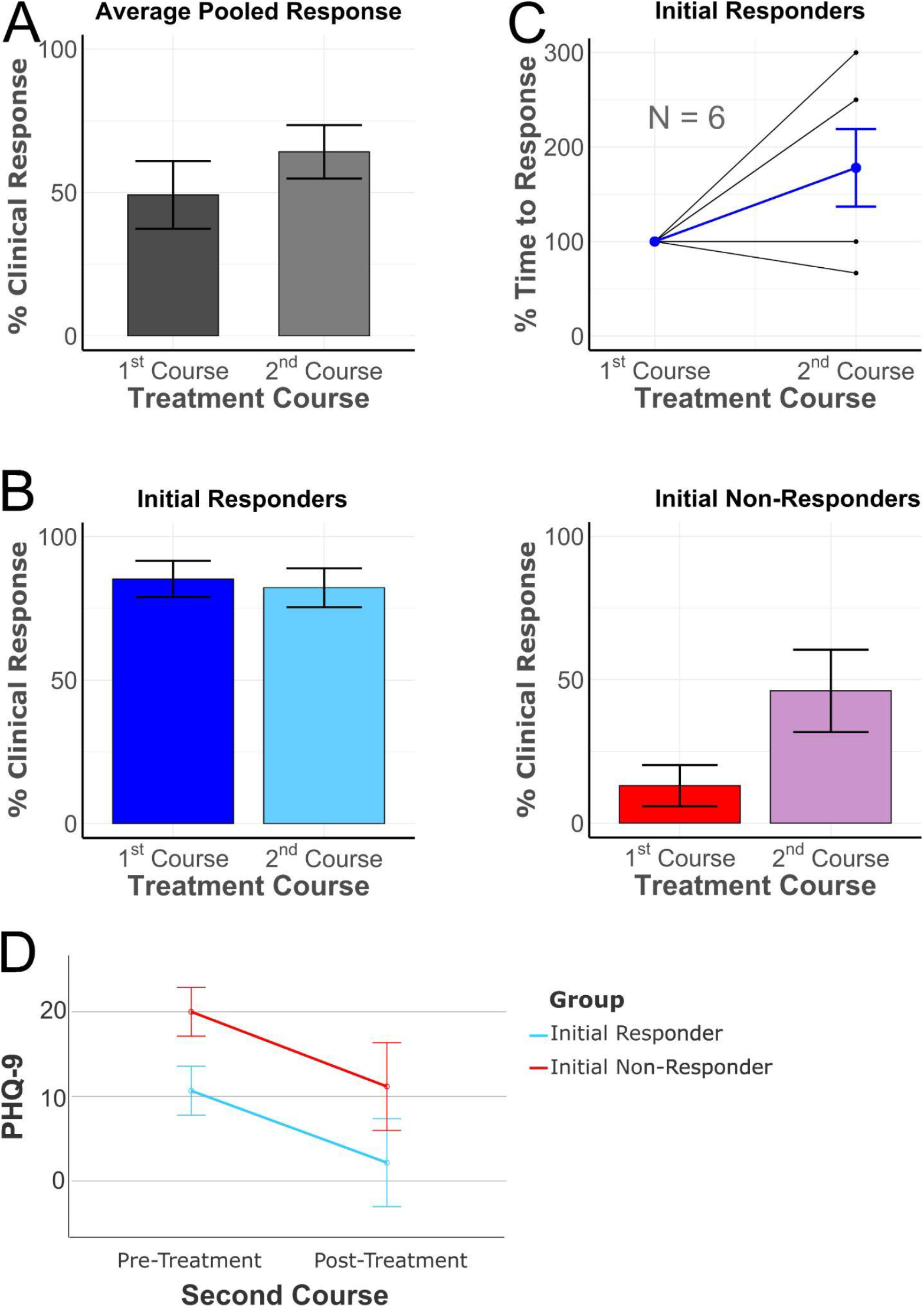
Relationship of initial clinical response to subsequent treatment response. A) Average clinical response of cohort (responders and nonresponders) in response to initial treatment course. B) Clinical response to second treatment course stratified by initial clinical response. C) Relative time to clinical response in second compared to initial treatment course. D) PHQ-9 scores in the second course between initial responders and non-responders. Error bars indicate standard error of the mean.

Of those demonstrating an initial clinical response (n = 6, >50% reduction in PHQ-9 scores), all six achieved clinical response in their second course of treatment (Fig 2A). On average initial responders demonstrated an 85.3% reduction of their depressive symptoms after their first course (Fig 1B, 2A, mean PHQ_initial_ = 13.21 ± 4.7., mean PHQ_final_ = 2.0 ± 2.0) and 81.3% after their second course (PHQ_initial_ = 10.7±3.4, PHQ_final_ = 2.3 ± 2.1). Individuals that demonstrated a clinical response to the initial treatment course presented on average 7 months following the end of the initial treatment course (7.0 ± 5.8 mo, nonresponders 16 ± 10.8 mo, p = 0.101) and with less baseline symptom severity (13.2 ± 4.7, nonresponders 20.5 ± 5.3, p=0.001) compared to those that did not respond to initial treatment. For initial treatment responders, the time it took to achieve clinical response during the second course varied widely, with 3 of 6 Veterans responding in the same amount of time or less than the time of the initial treatment, and the other three taking longer to respond during a second treatment (Fig 1C, normalized t(5) = 2.57, p = 0.1145; non-normalized t(5) = 2.57, p= 0.1579). Similarly, 3 of the 6 treatment responders that achieved symptom remission (PHQ-9 < 5) during the first treatment course attained remission in the same amount of time or more quickly during the second treatment course.

**Figure 2.**
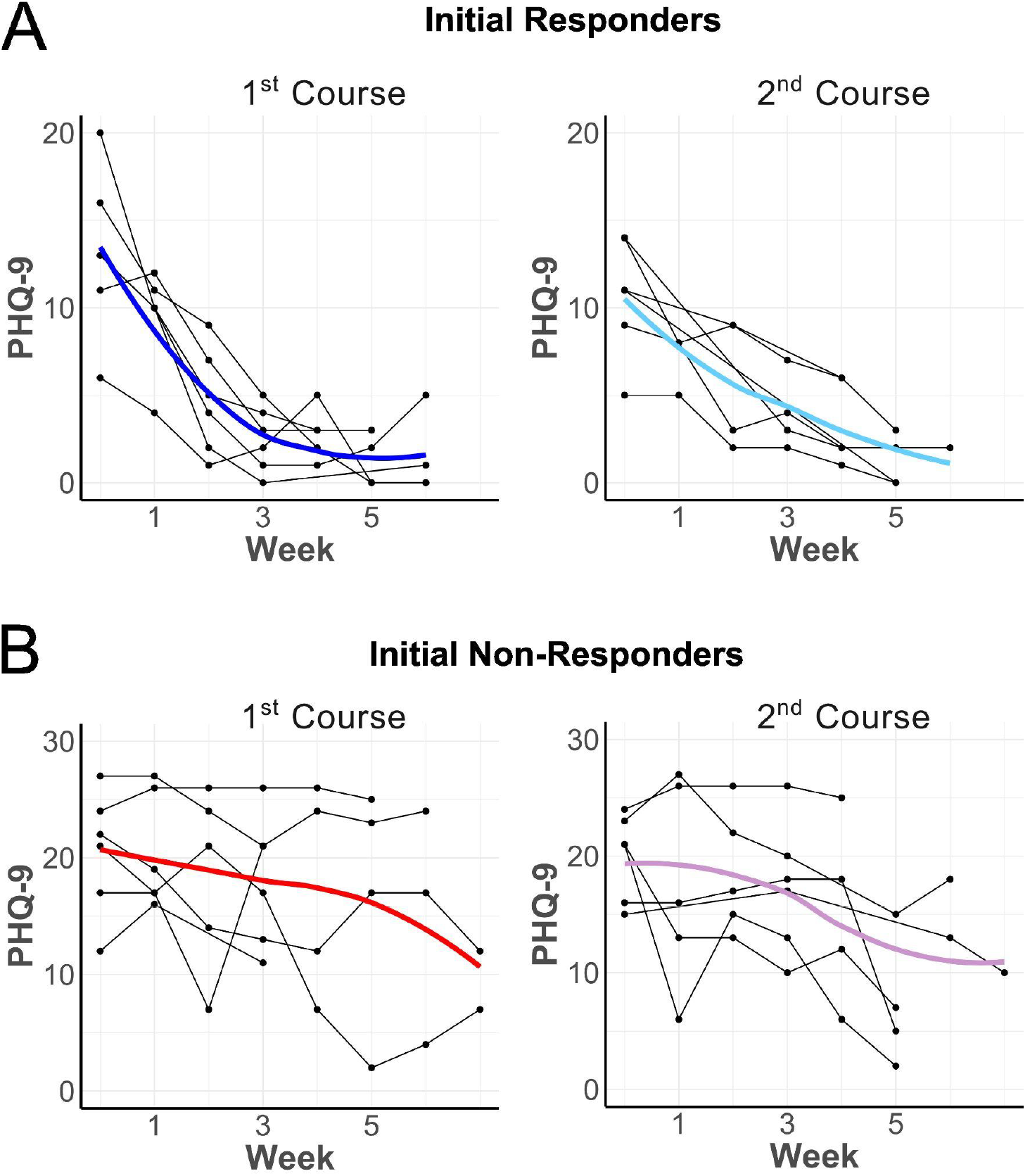
Additional TMS treatment courses demonstrated consistent benefit for initial treatment responders. Individual and group second course clinical responses for A) initial responders and B) initial non-responders. A) Note that 6 / 6 Veterans that demonstrated clinical response to the first course also responded in the second course. B) Initial non-responders demonstrated more variability, with half responding (n= 3 / 6) in a second course.

Initial non-responders (n=6) demonstrated a 13.1% reduction in depressive symptoms during that first course (Fig 1B, 2B, mean PHQ_initial_ = 20.5 ± 5.3., mean PHQ_final_ = 17.8 ± 6.2) and a 46% reduction after the second course (Fig 1B, 2B, mean PHQ_initial_ = 20.0 ± 43.7., mean PHQ_final_ = 11.2 ± 8.7). 3 of the 6 initial non-responders responded clinically to a second treatment course.

After demonstrating normality of the distributions of clinical scores in initial responders (Shapiro-Wilk test, N=6, initial course W=.906, p=.411, second course W=0.915, p=.469) and initial non-responders (N = 6, initial course W=.880, p=.269, second course W=.958, p=.802), a repeated measures ANOVA was performed to compare the percent clinical response in the second course between initial responders and non-responders. There was a statistically significant main effect on PHQ-9 score across the pre- and post-treatment time points in the second course (F(1,10) = 30.623, p < 0.001). Post hoc tests revealed PHQ-9 scores were significantly lower in the post-treatment time point (M = 6.67, SE = 1.83) than in the pre-treatment time point (M = 15.33, SE = 1.02, mean difference = 8.667, 95% CI [5.177, 12.156], p < 0.001). There was a significant interaction effect of initial treatment response on PHQ-9 scores in the pre- and post-treatment time points in the second course (F(1,10) = 13.204, p = 0.005, η2 = 0.569). Overall, PHQ-9 scores in the second course changed significantly within subjects, and varied significantly between subjects based on whether they responded in their initial treatment course (Fig 1D).

### Baseline depression severity and initial treatment response predicted symptoms over time during the second treatment course

We next asked how pre-treatment clinical features and initial clinical response predicted PHQ-9 scores across time during a second treatment course. A multiple linear regression model was run to predict PHQ-9 scores across time in the second course using pre-treatment PHQ-9 score, number of weeks into treatment, and initial treatment response as independent variables. Initial treatment response was calculated by subtracting post-treatment from pre-treatment PHQ-9 scores in the first course. Pre-treatment PHQ-9 score during the second course was positively correlated with PHQ-9 across all time points (Fig 3A; Estimate 0.66, 95% CI 0.42 to 0.91, p < 0.001). Time was negatively correlated with PHQ-9 (Fig 3B; Estimate -1.53, 95% CI -2.09 to -0.97, p < 0.001). Initial treatment response was found to be predictive of second-course PHQ-9 scores across time points (Fig 3C; Estimate -0.43, 95% CI -0.68 to -0.20, p < 0.001). Higher initial treatment response was associated with a larger decrease in PHQ-9 during the second course (Fig 3; adjusted R-squared = 0.723, F = 52.36, df = 3, 56; p <2.93*10^-16^). A similar approach was also attempted using the BSI-18. Linear regression was run to predict BSI-18 global severity index across time in the second course using baseline disease severity, time, and initial treatment response and was found to be highly predictive (Adjusted R-squared = 0.79, F=74.82, df = 3, 55, p = 2.2*10^-16^, Fig S1, Supplement). In summary, our findings indicate that baseline symptom severity, time, and initial treatment response are able to predict symptoms in a second TMS treatment

**Figure 3.**
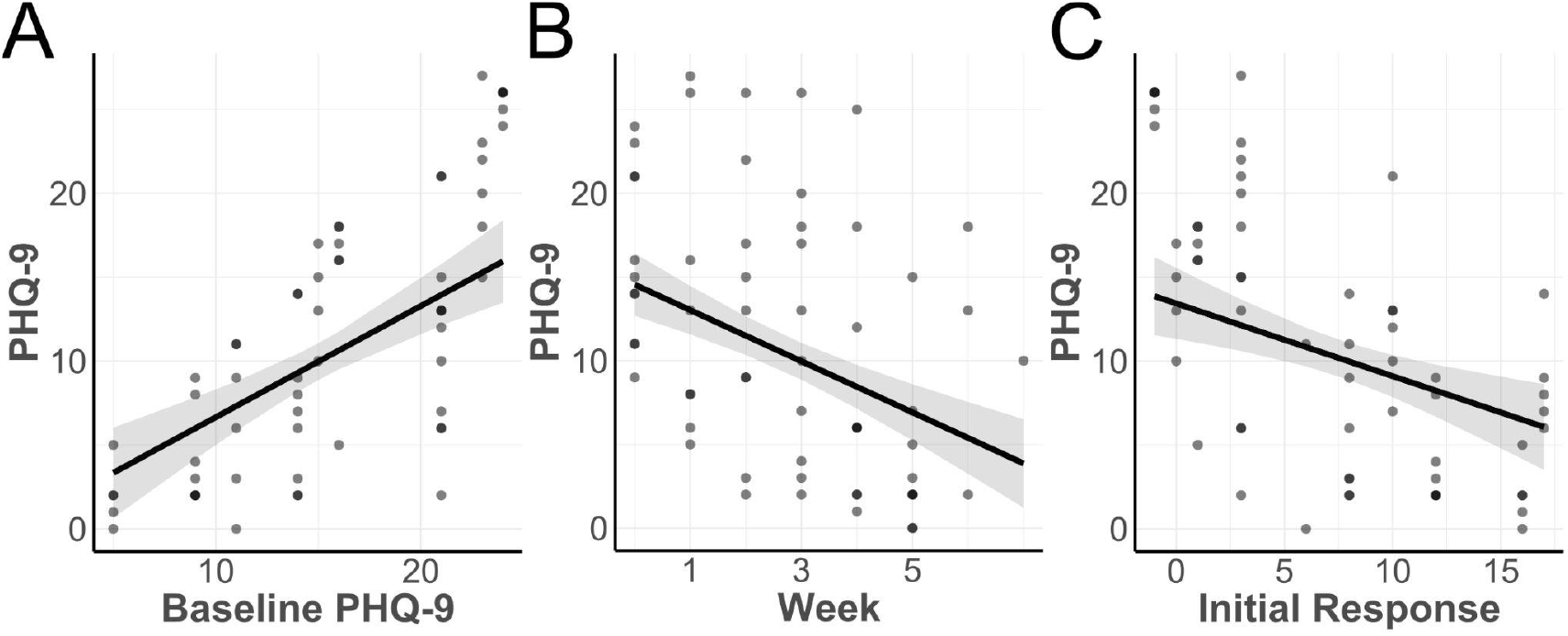
Multiple pre-treatment clinical features predict clinical response to a second treatment course. Multiple linear regression predicting PHQ-9 in a second course was calculated as a function of second course pre-treatment PHQ-9, time into the second course (weeks), and prior treatment response demonstrated in the initial course. Initial treatment response was calculated by finding the difference between initial treatment pre- and post-treatment PHQ-9 scores. These plots visualize the predictive contribution of each individual variable. The shaded region represents the 95% confidence interval. The black lines plot the correlation, with increasing magnitude of slope indicating a stronger relationship between the x-axis variable and PHQ-9 in this model. A) Baseline symptom severity prior to TMS treatment was directly correlated, indicating higher pre-treatment PHQ-9 scores were associated with higher scores later in the course. B) Time was negatively correlated, indicating PHQ-9 scores decreased through treatment. C) Initial treatment response was negatively correlated, indicating that a higher initial treatment response was associated with lower PHQ-9 scores in the second course.

### Patient response to treatment courses beyond the second

As an exploratory analysis, additional acute TMS courses beyond the second course were also examined (Fig S2, Supplement). Of the original 12 Veterans, three continued on for a subsequent third and fourth course, including two initial responders and one initial non-responder. The initial non-responder demonstrated a clinical response in their second course, supporting the recommendation for future treatments, if needed. This Veteran’s clinical response was sustained up to a fifth course. All initial responders continued to demonstrate clinical benefit to TMS treatment in all subsequent treatment courses.

## Discussion

Here we sought to investigate the clinical response in Veterans with TRD following repeated courses of TMS. This work supports the growing understanding that repeated courses of TMS are safe, well-tolerated, and effective in those that have demonstrated a clinical response to an initial treatment course. 100% (6 / 6) of Veterans who responded to their initial course of TMS also responded to their second course of treatment (Fig 1,2). In contrast, initial treatment non-responders demonstrated more variability, with half showing clinical benefit in a retrial. This differential clinical response to a second course of TMS was apparent despite confounders and barriers present in a real-world setting. Psychiatric comorbidity, adjunctive treatments, demographics, and environmental stressors were not controlled for in this sample. In summary, our findings show individuals who respond to an initial course of TMS in a real world setting are likely to respond to a second course.

We additionally identified pre-treatment clinical features that may predict response and thus be useful in planning a second treatment course. In a linear regression model, we were able to explain 72% of the variance in PHQ-9 by considering week into treatment, clinical response to an initial course, and pre-treatment depression severity (Fig 3). As it follows, a patient presenting with severe baseline depression and a poor initial response may require a longer time until potential treatment response in the second course. Similarly, a patient with more mild-to-moderate depression and robust initial response may demonstrate treatment response earlier. In our sample, initial treatment responders demonstrated lower baseline symptom severity (Table 2). Interestingly, the three initial non-responders who demonstrated benefit in their second course each had either reduced baseline symptom severity in the second course or were very close to meeting clinical response criteria in the initial course. Pre-treatment clinical features also predicted treatment response when using the same model structure for the BSI-18 (Fig S1, Supplement), indicating other symptoms not captured by the PHQ-9 may also be of value. The BSI-18 may be a helpful adjunct particularly in older Veterans with depression, anxiety, and tendency toward somatization (Gould et al., 2013).

It is important to examine the six Veterans who did not respond to their first trial of TMS. These Veterans were offered subsequent courses based on a clinician assessment of benefits versus risks despite not responding to initial treatment, as assessed by the PHQ-9. They demonstrated the highest pre-treatment PHQ-9 scores, consistent with prior work indicating less severe symptoms may be more predictive of TMS response (Grammer et al., 2015; Trevizol et al., 2020) (Table 2). Notably, the three youngest individuals in this group also reported the highest scores overall. Prior work has demonstrated that Veterans demonstrate unique cohort effects associated with different military-associated stressors (Gould et al., 2015; Kar, 2019). Younger Veterans active during the Global War on Terror (2001 - 2021) have demonstrated elevated levels of depression and anxiety compared to their counterparts. Two of these Veterans did not respond to a retrial, while the third, consistent with our model, demonstrated response following a near clinical response in a first course. In summary, higher baseline symptoms, later presentation, and younger age was associated with lack of clinical benefit in initial non-responder Veterans in their second course of TMS.

There are no current national standardized recommendations for repeated courses of TMS. Previous work noted significant variability in treatment protocols, with reliance on primarily open-label and case series studies (Wilson et al., 2022). At the very least, a single course of acute TMS for a depressive episode will be insufficient for the majority of people (Wilson et al., 2022). The lack of strict guidance, however, is not unusual for neuromodulation, as standardized guidelines for maintenance electroconvulsive therapy (ECT) also allow for clinician flexibility (Gill and Kellner, 2019). With TMS some clinics may follow a unique taper strategy, some to a target fixed frequency, others to gradually increasing intervals, while others adopt an “as needed” approach offering rescue treatments on signs of early relapse (Gill and Kellner, 2019). The American Psychiatric Association has also maintained the need to use the lowest dose and frequency necessary (American Psychiatric Association. Task Force on Electroconvulsive Therapy, 1990). These guiding principles help providers when selecting a program and evaluating risk versus benefit for patients with a spectrum of disease severity, treatment tolerability, and prior demonstrated response. FDA clearance for ECT offers similarly flexible language leaving discretion to the provider and notes that a significant proportion of patients may relapse. While FDA guidance for TMS offers text around acute treatment, preservation treatment has yet to be formally acknowledged. Evidence suggests that individuals who respond to TMS, as demonstrated in ECT, will likely require an additional course at some point.

While evidence supports the use of subsequent courses of TMS, financial factors limit patient accessibility. Repeated courses of TMS are not routinely covered by insurance and there are significant obstacles to obtaining coverage through insurance. The lack of financial reimbursement presents a barrier for patients, further limiting access as well as the ability to examine similar naturalistic designs outside of the Veteran health system (Wilson et al., 2022). The problem is a difficult one to overcome, as the lower quality of evidence to support financial coverage is further hindered by the lack of treatment access. As preservation TMS is not routinely prescribed or planned prior to an initial treatment course, access is likely influenced by individual patient motivation and demonstrated decompensation. As it currently stands, our group consisted of Veterans without significant work commitments and the benefit of VA-provided supports such as transportation, housing and financial subsidization. Having a rational approach to the financial aspects of preservation TMS will improve access and likely outcomes for those who have previously demonstrated a positive response.

There are several limitations to be considered in this analysis. Although the naturalistic design supports generalizability of the findings, the lack of rigorous controls allows for confounds. The sample size is small, limiting power. Second courses of TMS are not yet widely adopted in clinical practice but are offered in this clinic to those motivated Veterans who had demonstrated prior tolerability, which should be noted as a bias in this sample. Others were brought in with the advocacy of their referring provider due to ongoing refractory symptoms. Furthermore, Veterans varied in their baseline severity of symptoms prior to their second TMS course, with those more acute qualifying for a second course of TMS while those with milder scores on standard clinical surveys instead on the spectrum of relapse-prevention. Given the small sample size, these Veterans were examined together as a group in order to address the primary question; however, this may miss more subtle differences. Lastly, because this data were obtained from a consultation clinic, Veterans were assessed and offered repeated TMS courses in follow-up encounters 3- and 6-months post-treatment. The standardized approach does facilitate engagement but may not completely capture reemergence of symptoms past the 6-month time point.

Overall, these findings support the growing understanding that a second acute TMS course is safe, well-tolerated, and for those that respond in the initial course appear to provide clinical relief, at least in this small population. Larger powered studies are necessary to determine the efficacy of subsequent TMS treatment courses and to advocate for reimbursement by funding entities. Longitudinal designs will help determine the long-term safety and tolerability of repeated TMS courses. Regardless, this small retrospective study demonstrates the feasibility and utility in offering a second course of TMS for Veterans.

## Data Availability

All data produced in the present work are contained in the manuscript

## Acknowledgments

We are immensely grateful for the Veterans who received treatment in the clinic and are reported on here. Additionally, we thank the VA Office of Mental Health and Suicide Prevention for their support of the National VA TMS Program, through which data was collected. Lastly, this research is supported by the Department of Veterans Affairs Office of Academic Affiliations Advanced Fellowship Program in Mental Illness Research and Treatment, the Medical Research Service of the Veterans Affairs Palo Alto Health Care System, and the Department of Veterans Affairs Sierra Pacific Mental Illness, Research, Education, and Clinical Center (MIRECC).

## Supplemental Material

**Figure S1.**
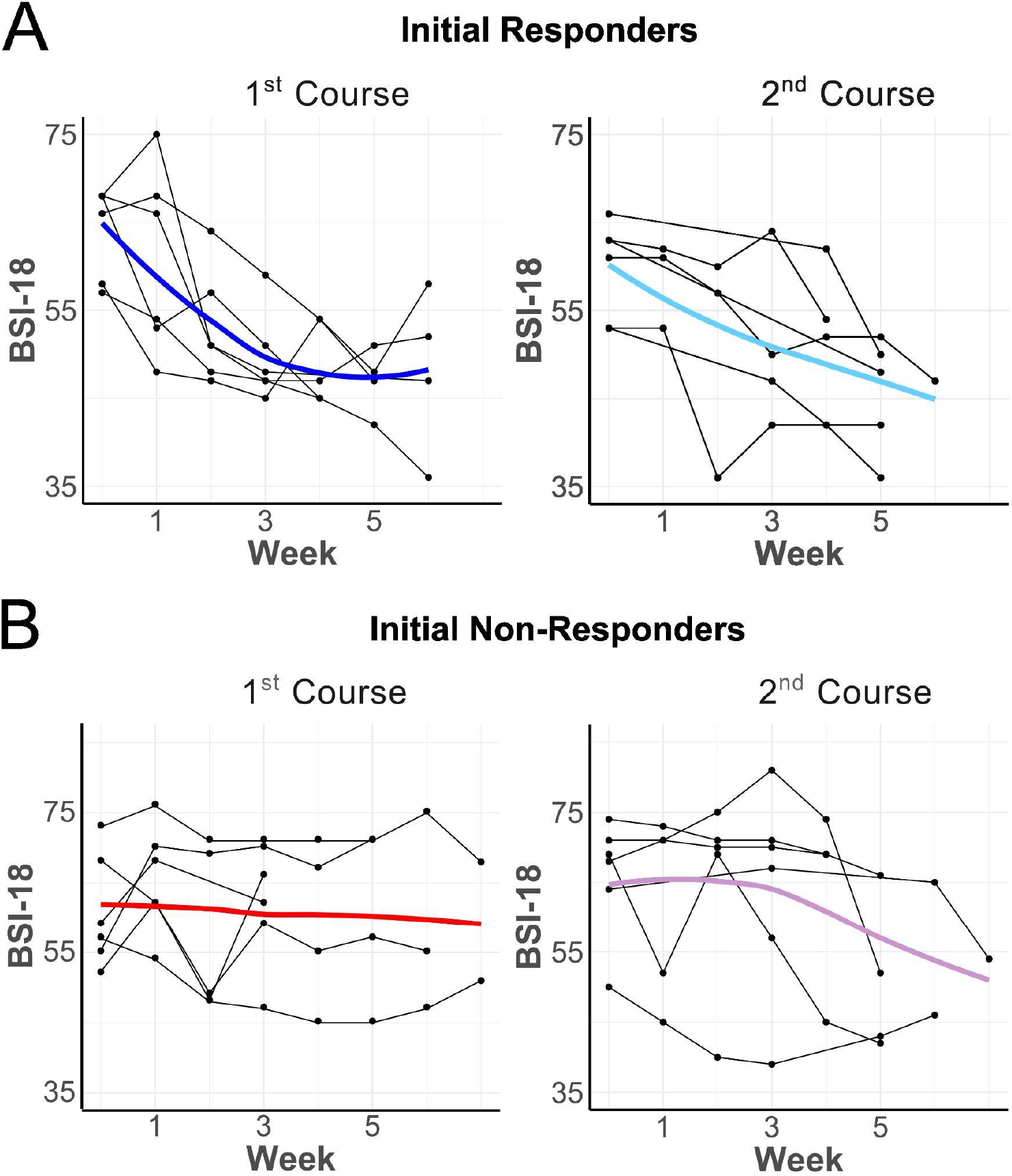
A) Initial responders (n = 6 / 12) demonstrated a similar pattern of response on the Beck Symptom Inventory, with a total 24.9% in the first and 28.1% in the second. B) Initial non-responders (n = 6 / 12) showed an average 6.2% reduction in the first course and 5.3% decrease in the second. The continued decline in symptoms over time noted in the figure is indicative of changes in longer treatment courses.

**Figure S2.**
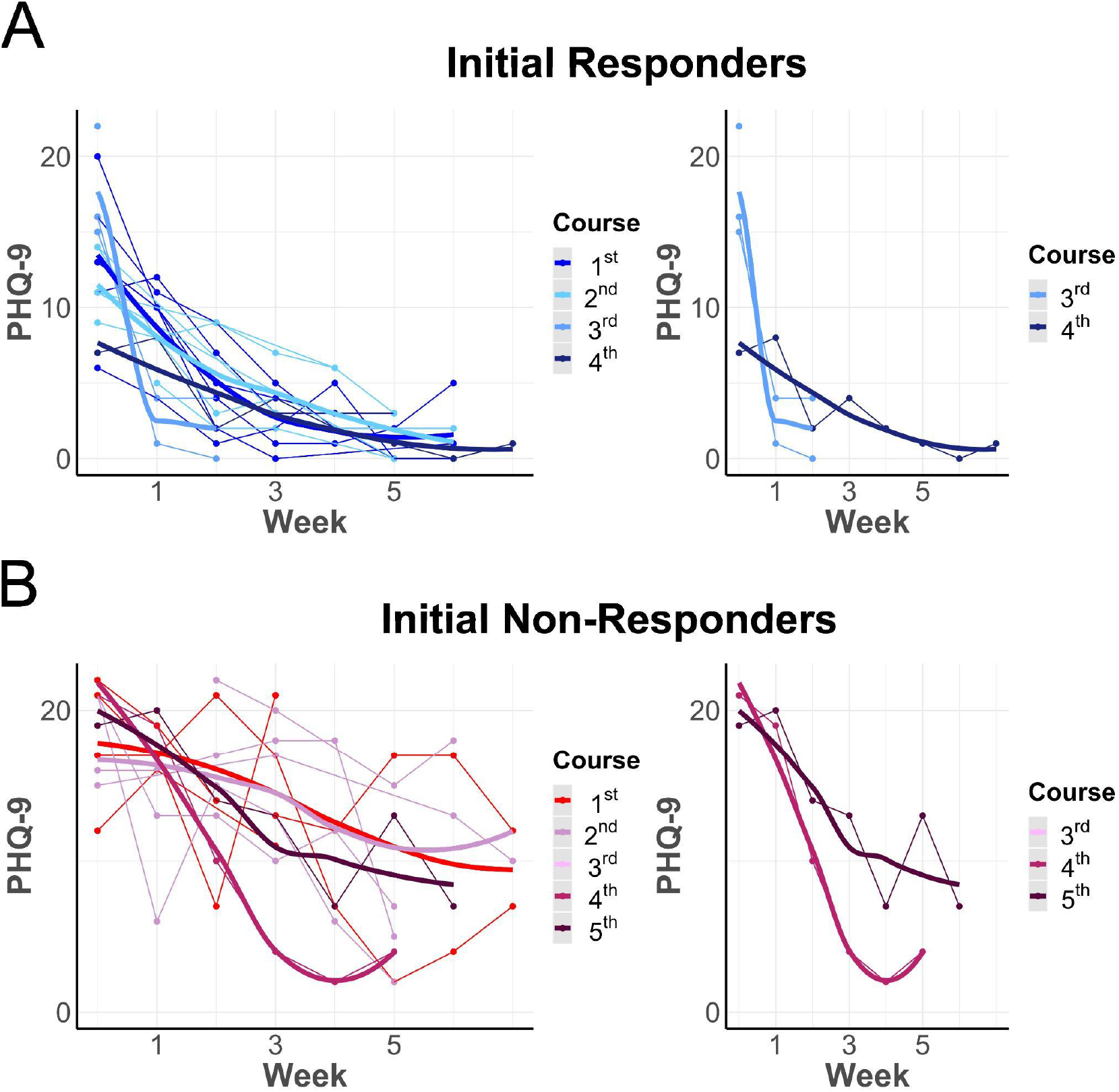
A) Initial Responders continued to demonstrate benefit in subsequent courses beyond the second course. Of the original 12 Veterans, 3 continued on for a subsequent third and fourth course, including two initial responders and one initial non-responder. B) One initial non-responder went on to demonstrate treatment response in the 2nd course, supporting the recommendation for future courses. This response was sustained up to the 5th course. Significance tests were not run in this exploratory analysis given the small sample of cases beyond the second course.

